# Occurrence of enteroparasites in vegetables commercialized in São Paulo, Brazil

**DOI:** 10.1101/2023.01.30.23285185

**Authors:** Thais Boccia, Andrea Bento Lopes

**Affiliations:** Department of General Surgery, Boston Children’s Hospital, Massachusetss, USA; Universidade Presbiteriana Mackenzie, São Paulo, Brazil

## Abstract

Parasitic diseases constitute an important public health problem. Their transmission can occur by ingestion of water contaminated with human feces and/or parasitized animals, and by poorly washed fruits and vegetables. Considering the scarcity of national studies reporting the degree of contamination of vegetables, the present study investigated and compared the contamination of enteroparasites in 32 samples of each of the types: lettuce (*Lactuca sativa*) of the smooth and curly variety and watercress (*Nasturtium officinale*), commercialized in the city of São Paulo. For this purpose, the samples were washed with distilled water. The product of the washings was decanted in conical sedimentation vessels for 24 hours by the spontaneous sedimentation method, and the sediment was analyzed under an optical microscope. The results showed that of the 96 samples analyzed, 78 (82%) presented some parasitic form. Vegetables purchased from supermarkets and street markets showed a contamination rate of 80% (48/60) and 83% (30/36), respectively. The parasitic forms found were: 42% *Ancylostoma sp* larvae, *15*.*94% Ancylostoma sp* eggs, 7.24% *Ascaris sp* eggs, 8.69% *Enterobius sp* eggs, 8.69 % *Strongyloides sp* larvae, 7.24% *Hymenolepis sp* eggs, 5.79% *Taenia sp* eggs and 4.34% *Entamoeba sp* cysts. These results demonstrate the need to develop a more efficient health education program aimed at horticulturists and vegetable handlers, with the intention of preventing the occurrence of these parasites, and further contamination of the population.

## Introduction

Infections by helminths and endoparasites constitute a serious public health problem, as, according to the World Health Organization, they are among the most frequently aggravating infectious diseases in the world (Health Organization, 1987; Jourdan et al., 2018; Li et al., 2020). In Brazil, the prevalence of these infections in some locations is still considered high (Ferreira et al., 2000; Leite et al., n.d.; Neres et al., 2011). Enteroparasitic infections can affect the nutritional balance and cause significant complications, especially in school-age children (Nussenzveig et al., n.d.; Prado et al., 2001), because they have an infectious ecosystem, with a passive oral or active cutaneous infection mechanism (Mesquita et al., 1999). The significant morbidity and opportunity for infection by one or several intestinal parasites is worldwide, due to the widespread of these agents and the ease with which they are transmitted (Jourdan et al., 2018; Li et al., 2020).

Developing countries have high levels of demographic growth, which the health structure cannot keep up with, providing conditions for the perpetuation of the biological cycle of parasitic infections (Vieira, n.d.). In underdeveloped countries, intestinal parasites reach infection rates of up to 90%, with a significant increase in frequency as the socioeconomic level declines. In Brazil, intestinal parasites are among the main public health problems (Almeida et al., 1991). The infection rate of some helminths in the state of São Paulo is still a serious public health problem, especially in the metropolitan region of the capital (Waldman & Chieffi, 1989), and control measures against helminths and intestinal protozoa have been relatively insufficient (Neves, n.d.).

The presence of parasitic forms in the soil, eliminated by human or animal defecation (Oliveira & Germano, 1992; Rey, 1991), or in the hands of food handlers (Neves, n.d.), mechanical vectors such as flies and cockroaches (Moraes & Leite, n.d.) and the water used to irrigate vegetables are important sources for the dissemination of intestinal parasites (Faria et al., 2014; Guilherme et al., 1999; Guimarães et al., 2003). Vegetables being cultivated in places with low sanitary conditions act as a vehicle for transmission of these parasitic forms (Chitarra, 2000; de Almeida Marzochi, 1970). Vegetables that grow in polluted soils can carry eggs of helminth parasites of humans, mainly *Ascaris sp, Trichuris sp, Taenia sp, Hymenolepis sp, Ancylostoma sp* eggs and larvae, *Strongyloides sp*, and also protozoan cysts (Hernandes et al., n.d.).

In Brazil, there are few studies that establish the rate of contamination of vegetables by intestinal helminths. (Gelli et al., 1979), showed that samples of vegetables sold in the city of São Paulo were positive for eggs and/or larvae of *Ancylostoma sp* (59.3%), and *Strongyloides sp* (5.3%). Contamination by these organisms was observed in 84.6% of arugula, 78.3% of watercress, 53.3% of endive, and 32.4% of lettuce samples.

Hernandes et al, 1981 found *Ascaris sp* and hookworm eggs in vegetables from 12 vegetable gardens in the municipality of Biritiba Mirim, SP, one of the main vegetable producers in the state. Likewise, (Mota et al., n.d.), in Curitiba, and (Miche & Morganti, n.d.), in São Paulo, analyzed 154 and 68 samples of vegetables respectively, detected the occurrence of high levels of intestinal parasites. In Paraná, in the municipality of Maringá, (Guilherme et al., 1999), analyzed 144 samples of vegetables and found watercress as the most contaminated (100%) followed by mimosa lettuce (25%) and plain lettuce (21%). (Guimarães et al., 2003), analyzed 120 samples of lettuce acquired in supermarkets and street markets and observed nematode larvae (47.5%), *Entamoeba sp* cysts (5%), and insects (34.2%). Bacteriological analysis of 81 water samples from 44 rural properties, used, among other purposes for crop irrigation, showed that almost all of the springs were contaminated by fecal coliforms.

More recently, (Rocha et al., 2021) analyzed lettuce samples in the city of Goiânia, Brazil, and found 45% of contamination with helminths eggs, protozoan cysts and oocysts. The more frequent parasitic forms were eggs of *Ancylostomatidae, Strongyloididae, Ascarididae* and *Taeniidae*, or oocysts of *Eimeriidae, Cystoisospora sp*. and *Toxocara sp*. Also, (Mendonça Ambrozim et al., 2017) analyzed 120 samples from the state of Espírito Santo, Brazil, and 59% had one or more parasitic contaminants, and in lettuce the contamination rate was 78.3%, and in parsley 40%. (Machado et al., 2018) showed the presence of total coliforms in vegetables, and also the contamination with enteroparasites, mainly *Entamoeba sp*., *Balantidium coli, Strongyloides sp*., *Ascaris sp*., *Enterobius vermicularis*, and *Ancylostomidae* in vegetables comercialized in street markets from all regions of Brazil.

Techniques for parasitological examination of food are still poorly developed. Most procedures aim at the concentration of eggs and larvae in the samples, through techniques such as spontaneous sedimentation, simple centrifugation, simple centrifugation associated with the centrifuge, flotation, and ultracentrifugation. The fact that the high rate of consumption of raw vegetables by the population and the importance of transmission of enteroparasites is inversely proportional to the number of studies found in the literature that inform the degree of contamination, led us to carry out this work.

## Methods

### Samples

In the period between March and September 2005, a total of 96 vegetables were purchased, among them: lettuce (*Lactuca sativa*) (n=64) of the smooth and curly varieties and watercress (*Nasturtium officinale*) (n=32), from supermarkets and street fairs in the city of São Paulo. Each lettuce sample consisted of a stalk, regardless of weight or size, while the watercress samples consisted of a bunch.

### Procedure

The vegetables were stored in sterile plastic bags of approximately 5 L, and refrigerated in a refrigerator at 4ºC until the moment of washing. For washing, 200 mL of distilled water at room temperature were poured into a new sterile plastic bag for a more superficial washing. This bag was slowly shaken so that everything adhered to the leaves was removed by the force exerted by the water. The water contained in the plastic bag was filtered through filter paper and poured into a conical-bottomed cup for sedimentation. After this initial process, the vegetable was defoliated and each leaf was washed with another 200 ml of distilled water. The washing of the leaves was manual with the concern to wash all the recesses on both sides of the sheet.

The water contained in the cups was left untouched for a maximum of 24 hours. The sediment accumulated was removed with the aid of a pipette and stored in small flasks with the addition of 2 mL of Merthiolate-Iodine-Formaldehyde (MIF) so that the forms present in this sediment were preserved until the moment of analysis.

For analysis of the sediment, a glass slide was prepared with the addition of a drop of Lugol, and the reading was performed under an optical microscope at 400X magnification.

## Results and discussion

Of the 96 samples of smooth or curly lettuce, and watercress acquired randomly, 78 (81.2%) contained some kind of parasitic form. Dividing by origin, we see that 80% of the samples collected from supermarkets and 83% of the samples from street markets were positive for some kind of parasitic form. In samples purchased in supermarkets, we observed that parasitic forms were present and the three varieties of vegetables showed the same positivity rate (80%) (Table 1).

**Table 1:** Frequency of forms of intestinal parasites in vegetables acquired in supermarkets and street markets in the city of São Paulo, Brazil. A total of 20 samples of each vegetable were collected from supermarkets, and 12 samples from street markets.

| Sample | Supermarket | Steet market |
| --- | --- | --- |
|  | Positive (%) | Positive (%) |
| Watercress | 16 (80%) | 10 (83%) |
| Smooth Lettuce | 16 (80%) | 11 (91%) |
| Curled Lettuce | 16 (80%) | 9 (75%) |
| Total | 48 (80%) | 30 (83%) |

The analysis of the parasitic forms observed in the samples of watercress, smooth and curled lettuce showed a predominance of *Ancylostoma sp* larvae in the three types of vegetables, being more frequent in watercress (Figure 1). Regarding *Ancylostoma sp* larvae and eggs, we observed a similar frequency between samples of smooth and curled lettuce, and watercress, both in supermarkets and in street markets. Only in supermarkets, larvae of *Strongyloides sp*, eggs of *Hymenolepis sp, Taenia sp*, and *Entamoeba sp* were observed in all types of vegetables, with the exception of the last two which could not be observed in curled lettuce. Both *Ascaris sp* and *Enterobius sp* eggs were observed in vegetable samples from supermarkets. In curled lettuce samples, we did not observe forms of these helminths in samples purchased from street markets (Figure 1).

**Figure 1:**
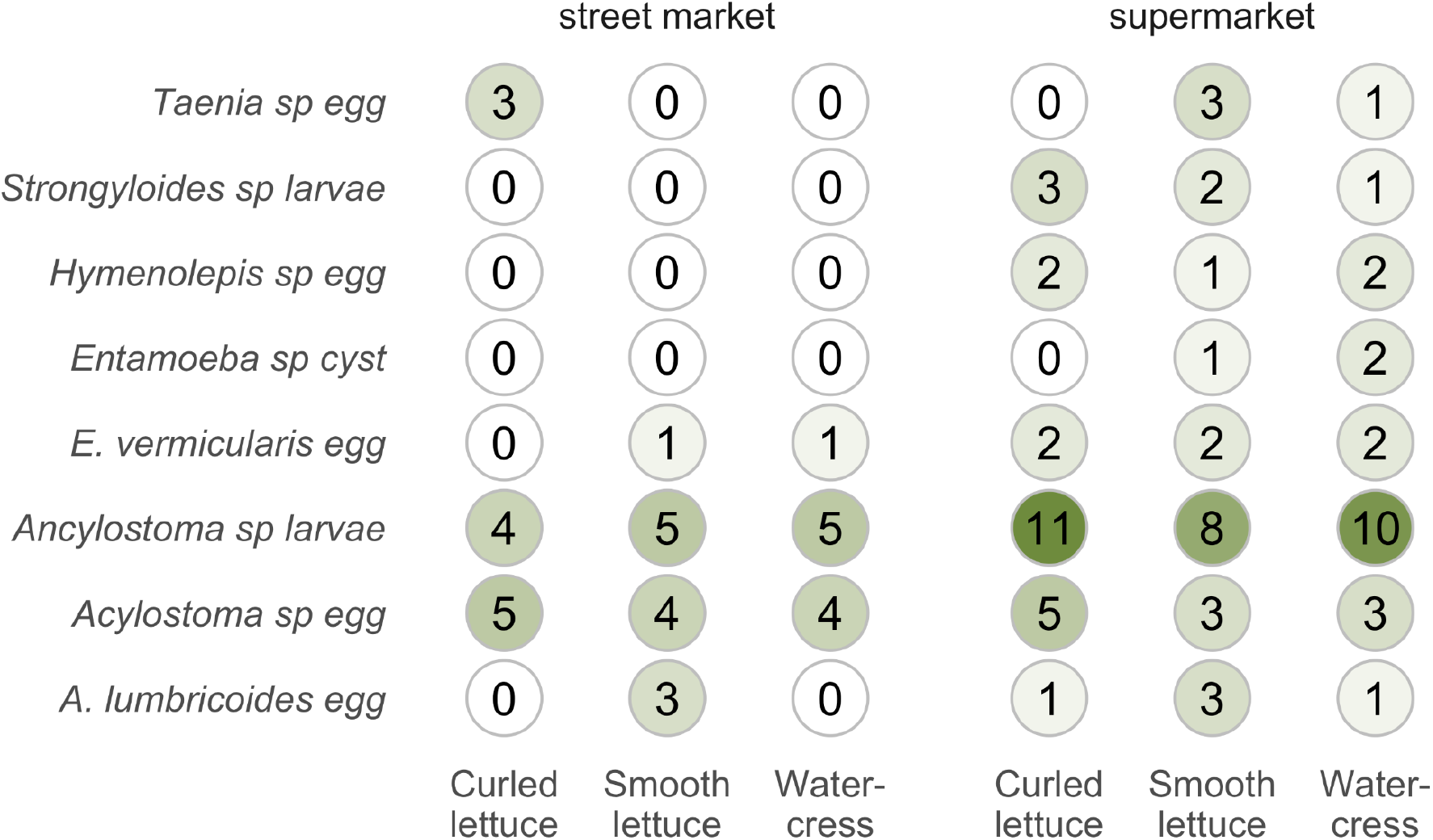
Distribution of the parasitic forms found in vegetables commercialized in supermarkets and street markets in the city of São Paulo, Brazil. A total of 20 samples of each vegetable were collected from supermarkets, and 12 samples from street markets.

The results obtained in the present work showed high percentages of contamination by parasitic forms in all the analyzed vegetable varieties, which suggest hygienically inadequate cultivation conditions and practices (Rude et al., 1984). These results are not in agreement with those observed by Oliveira and Germano 1992 (Oliveira & Germano, 1992), who found a percentage of 66%, 34%, and 32% of positivity in the samples of watercress, curled and smooth lettuce respectively, commercialized in the city of São Paulo.

The rate of parasitic contamination of vegetables sold in supermarkets in the city of São Paulo in this study was higher than 80%, a relatively high number when compared to the work by (Silva et al., 1995), who found 21.4% of contamination in samples from supermarkets sold in Rio de Janeiro. Also, different results were observed by (Mesquita et al., 1999), when analyzing vegetables sold in the cities of Niterói and Rio de Janeiro, in which only 6.2% of the studied samples presented some type of parasitic structure.

As the acquisition of the samples was made in a period of less rainfall, from March to September, the irrigation of the vegetables is done artificially (Oliveira & Germano, 1992), thus it can contaminate the vegetables since manipulated water can be contaminated. Perhaps if the collection was carried out in rainy seasons, the positivity rates could be lower (Oliveira & Germano, 1992). In addition to this artificial irrigation, the presence of parasitic forms in samples of smooth lettuce could be explained by the lack of hygiene during planting, as according to (Cristóvão et al., n.d.) there is greater care in the cultivation of lettuces, as they are sold in greater quantities. Once again, our results do not suggest this trend, since the smooth lettuce samples are still heavily contaminated.

The physical structure of the vegetables can contribute to the occurrence of different levels of contamination (Gelli et al., 1979). Both Oliveira & Germano, 1992 and Gelli et al, 1979 observed a higher frequency of parasitic forms in watercress samples, while Guilherme et al, 1999, comparing smooth and curled lettuce, observed that the former was more contaminated, suggesting the cultivation conditions and not only to the anatomical structure of the plant are influencing the contamination (Gelli et al., 1979; Guilherme et al., 1999; Oliveira & Germano, 1992).

The presence of *Ascaris sp* eggs in the samples may also be related to the fact that this specie do not need an intermediate host to develop their larval forms, only the presence of a favorable soil is enough, in addition to having a greater adhesion to the leaves of the vegetables due to the eggshell morphology (Coelho et al., 2001). *Strongyloides sp* larvae and *Hymenolepis sp* eggs are most commonly found in moist soils, as their forms are less resistant to dryness, in addition to indicating fecal contamination of human origin, since they present species that parasitize humans (Oliveira & Germano, 1992).

Samples contaminated with *Taenia sp* eggs, 3%, 9% (Figure 1) for watercress and smooth lettuce respectively, can be an aggravating factor for the appearance of human cysticercosis by the ingestion of eggs from *Taenia solium*. As the eggs released by the hosts present great resistance, the transmission of human cysticercosis by vegetables has been reported (de Almeida Marzochi, 1970).

Surprisingly, we observed eggs of *Enterobius sp* in samples of smooth lettuce and watercress, both from supermarkets (Figure 1). These eggs are not released in the host’s intestine, but in the perianal region, making it difficult to find them in human or animal feces samples, suggesting a contamination in the manipulation steps, not in the fertilizers (Neves, n.d.).

The presence of *Entamoeba sp* cysts in the samples indicates fecal contamination of human and/or animal origin. Although some species are non-pathogenic, they still present considerable levels of occurrence in the population of the metropolitan region of São Paulo (Oliveira & Germano, 1992). In addition, asymptomatic patients in rural or urban areas continue to be the major sources of transmission. However, the protozoan cysts found in the samples could not have their species identified, because it was not possible to observe the number of nuclei present in the cytoplasm. The larvae and eggs of *Ancylostoma sp*, the eggs of *Ascaris sp* and the larvae of *Strongyloides sp* were the most prevalent parasitic forms, which can be explained by the fact that they can survive for a longer time in the soil.

Considering the magnitude of the results obtained in this work, health surveillance activities should be concentrated on the production of vegetables and laboratory monitoring of artificial water intended for irrigation.

## Data Availability

All data produced in the present study are available upon reasonable request to the authors.

## Acknowledgements

The authors thank Mackenzie Presbyterian University for assisting in the development of the research.

